# Predicting successes and failures of clinical trials with an ensemble LS-SVR

**DOI:** 10.1101/2020.02.05.20020636

**Authors:** Zhen-Yu Hong, Jooyong Shim, Woo Chan Son, Changha Hwang

## Abstract

For a variety of reasons, most drug candidates cannot eventually pass the drug approval process. Thus, developing reliable methods for predicting clinical trial outcomes of drug candidates is crucial in improving the success rate of drug discovery and development. In this study, we propose an ensemble classifier based on weighted least squares support vector regression (LS-SVR) for predicting successes and failures of clinical trials. The efficacy of the proposed ensemble classifier is demonstrated through an experimental study on PrOCTOR dataset, which consists of informative chemical features of the drugs and target-based features. Comparing random forest and other models, the proposed ensemble classifier obtains the highest value for the area under the receiver operator curve (AUC). The results of this study demonstrate that the proposed ensemble classifier can be used to effectively predict drug approvals.

## 1 Introduction

In the analysis of the drug development costs of 98 companies in the past 10 years, the average cost of each drug developed and approved by a single-drug company was 350 million US dollars [1]. Although the extensive efforts and resources invested in identifying and elaborately designing new compounds as well as the systematic examination of all steps in the early development, even the most promising high-value compounds often fail in clinical trials. The probability of launch from entry to phase I has also remained below 10% [2]. An analysis of the causes of clinical failure in 2016-2018 shows that these are largely unchanged over the past 3 years [3]: 79% were due to safety or effectiveness; 1% were attributable to operational or technical shortcomings; 13% were the result of strategic realignment; and 7% were for commercial reasons [4, 5, 6]. It is one of the main challenges of the pharmaceutical industry to find innovative solutions for reducing the clinical failure rates, because failure in later clinical trials will lead to huge losses. At present, it is general practice to apply a variety of computational modeling approaches and simulation tools to upgrade and speed up drug discovery, design and other steps in the early development phase [7]. Moreover, the recently adopted computational-based strategy is to develop a set of criteria that can be applied to predict the results of clinical trials before they begin. A major focus of the criteria consists in the ability to identify compounds with adverse toxicity properties.

In fact, the prediction of successes and failures of drug candidates in clinical trials is a binary classification problem. In addition, this is imbalanced data classification problem, which is inevitable in this area. The group of data that has a larger number of examples is called the majority class or negative class, whereas the group of data that has a smaller number of examples is called the minority class or positive class. Most of the data in biometrics as sell as clinical trials are usually imbalanced and have high-dimensional multimodal feature. This seriously affects the classification performance of the model. [8] lately developed a classifier named PrOCTOR to deal with clinal drug toxicity on the basis of random forests that not only considers the bioavailability related properties of the drugs, but also target-related properties. The properties include established informative chemical features of the drugs with target-based features and tissue selectivity. In their study, the authors proved that some of these properties, even when considered separately, have a significant discriminative power. In addition, employing a feature importance analysis, they have exhibited that both the target-based and chemical features contribute to effective classification. In this paper, we develop an ensemble classifier based on weighted LS-SVR for bimodal imbalanced data, and then compare this model with random forest model on almost the same dataset as PrOCTOR dataset used in [8]. This dataset was provided by the corresponding author of [8].

Developing an effective classification method for imbalanced and multimodal data is a challenging task. The rest of this paper is organized as follows. In Section 2 we review the standard LS-SVR and study variants of LS-SVR for bimodal data. In Section 3 we study variants of LS-SVR for imbalanced data. Section 4 and section 5 present experimental study and conclusion, respectively.

## 2 LS-SVRs for bimodal data

Least squares support vector machine (LS-SVM), which has been successfully applied to a number of real problems of classification and function estimation, is least squares version of SVM [9] and was proposed by [10]. LS-SVM has been proved to be a very appealing and promising method. There are some strong points of LS-SVM. One is that LS-SVM uses the linear equation which is simple to solve and good for computational time saving. Another is that LS-SVM classification is actually equivalent to LS-SVM regression in binary classification case [11]. For brevity, LS-SVM regression is called LS-SVR. For this reason, we will develop classification method for bimodal data using LS-SVR technique.

### 2.1 LS-SVR

Given a training data set 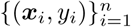 with each input ***x***_*i*_ and corresponding response *y*_*i*_, LS-SVR optimization problem in primal weight space is given as follows:

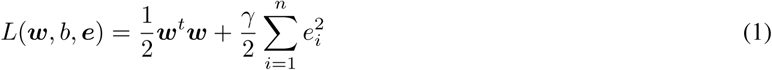

subject to equality constraints

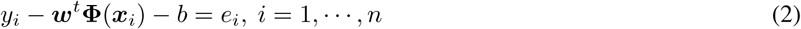

with a penalty parameter *γ* > 0, 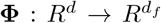 a function which maps the input space into a higher dimensional (possibly infinite dimensional) feature space, weight vector 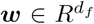 in primal weight space, error variables *e*_*i*_ ∈*R* and bias term *b*. To find minimizers of the objective function, we can construct the Lagrangian function as follows,

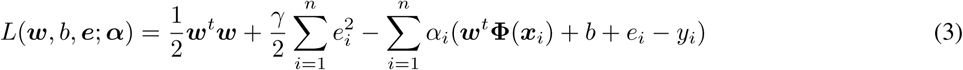

where *α*_*i*_’s are the Lagrange multipliers. Then, the conditions for optimality are given by

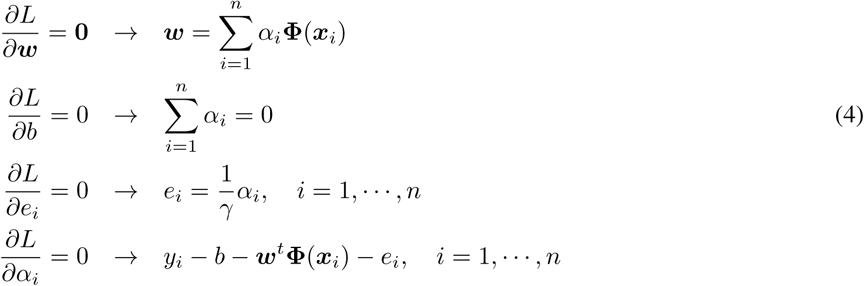

The estimation of parameters with equations in the above conditions requires the computations of inner products **Φ**(***x***_*i*_)^*t*^**Φ**(***x***_*j*_) in a potentially higher dimensional feature space. Under certain conditions these demanding computations can be reduced significantly by introducing a kernel function *K* such that **Φ**(***x***_*i*_)^*t*^**Φ**(***x***_*j*_) = *K*(***x***_*i*_, ***x***_*j*_) [15]. Possible kernel functions are linear kernel and radial basis function (RBF). The linear and RBF kernel are the most frequently used kernel function for the linear and the nonlinear case, respectively. After eliminating *e*_*i*_ and ***w***, we can obtain the optimal values of (***α***, *b*) from the following linear equations

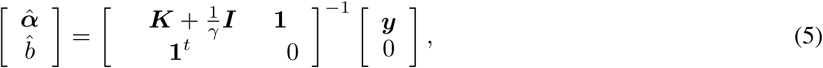

where ***K*** = {*K*_*ij*_} is the kernel matrix with *K*_*ij*_ = *K*(***x***_*i*_, ***x***_*j*_), *i, j* = 1,…, *n*.

Finally, for a given ***x***_*t*_ the predicted value of response is given as

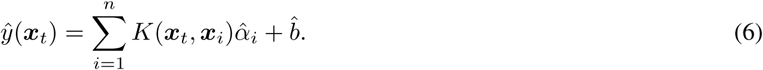

In particular, for the given training data set, we obtain

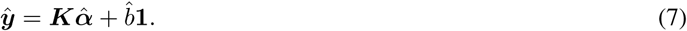

The functional structures of LS-SVR is characterized by the hyperparameters such as penalty parameter and kernel parameter. To choose optimal values of hyperparameters of the model we define a leave-one-out cross validation (LOOCV) function as follows:

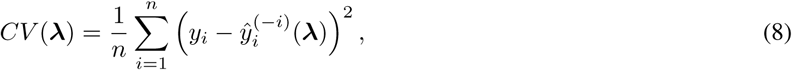

where ***λ*** = {*σ, γ*} is the set of hyperparameters and 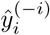 is the predicted value of *y*_*i*_ obtained from the data without *i*th observation. Since for each candidate of hyper-parameters, 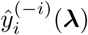 for *i* = 1, …, *n* should be evaluated, selecting parameters using CV function is computationally formidable. Generalized cross validation (GCV) function is obtained by applying the leaving-out-one lemma [12] as follows:

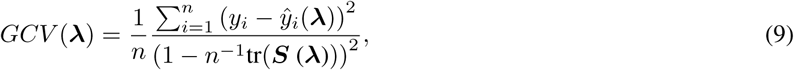

where 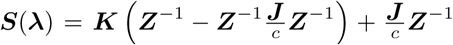. Here 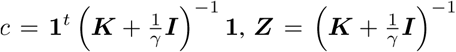 and ***J*** is a square matrix with all elements equal to 1. See for details [13].

### 2.2 Bimodal LS-SVR

Combining two different types of data for improving performance seems intuitively appealing task. We now devise LS-SVR model for bimodal data. For simplicity, it is named bLS-SVR. For bLS-SVR we consider two separate feature spaces, each of which is the molecular property related feature space and the target-based property related feature space. Then the optimization problem in primal weight space is written as follows:

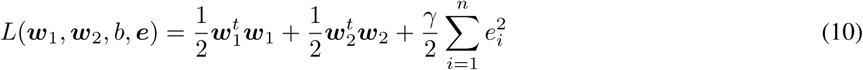

subject to equality constraints

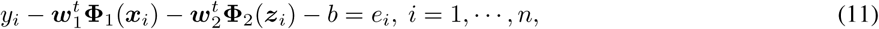

where ***x***_*i*_ and ***z***_*i*_ are the molecular property related input and the target-based property related input, respectively. To find minimizers of the objective function, we can construct the Lagrangian function as follows,

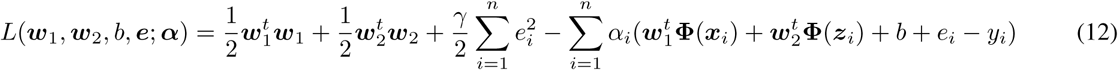

where *α*_*i*_’s are the Lagrange multipliers. The optimal values of (***α***, *b*) from the following linear equations

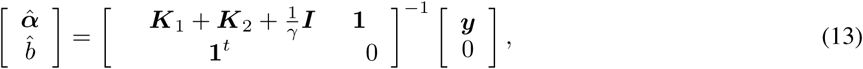

where ***K***_1_ and ***K***_2_ are the kernel matrices with elements *K*_1_(***x***_*i*_, ***x***_*j*_) and *K*_2_(***z***_*i*_, ***z***_*j*_), respectively.

Thus, for a given (***x***_*t*_, ***z***_*t*_) the predicted value of response is given as

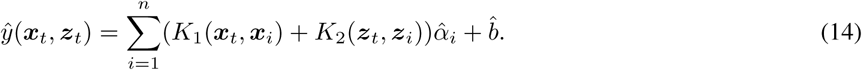

### 2.3 Stacked LS-SVR

To devise an LS-SVR reflecting the attribute of bimodal data, we borrow the idea of stacking which is an ensemble technique. For simplicity, it is named sLS-SVR. Because there two separate feature sets, we consider the ensemble algorithm to obtain better performance as follows:

1. Train LS-SVR using 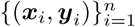 with the optimal values of hyperparameters and obtain *ŷ*(***x***_*i*_) and *ŷ*(***x***_*t*_) for given ***x***_*t*_.
2. Train LS-SVR using 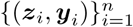 with the optimal values of hyperparameters and obtain *ŷ*(***z***_*i*_) and *ŷ*(***z***_*t*_) for given ***z***_*t*_.
3. Find regression coefficients *β*_0_, *β*_1_, *β*_2_ from the linear regression model with the input (*ŷ*(***x***_*i*_), *ŷ*(***z***_*i*_)) and the corresponding output *y*_*i*_.
4. Obtain the predicted value *ŷ*(***x***_*t*_, ***z***_*t*_) = *β*_0_ + *β*_1_*ŷ*(***x***_*t*_)*β*_1_*ŷ*(***z***_*t*_).

## 3 LS-SVR for imbalanced data

Since the ratio of approved drugs to those that failed for toxicity in clinical trials is imbalanced, the associated dataset is highly imbalanced. Thus, we now study some variants of LS-SVR for imbalanced data.

### 3.1 zLS-SVR

In SVM [9] the classification decision function can be written as follows:

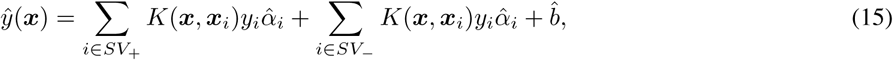

where *y*_*i*_ = 0 or 1, *SV*_+_ and *SV*_−_ are the index sets of support vectors corresponding to positive and negative *y*_*i*_’s, respectively. To reduce the bias of a learned SVM to majority class for imbalanced data, [14] introduced a multiplicative weight *z*, associated with each of the positive class support vectors. Under the assumption that minority class is considered as positive class, they reformulated SVM classification decision function as follows:

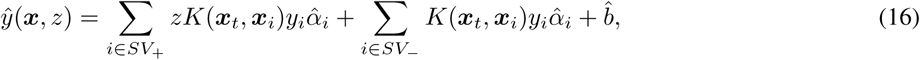

This can be regarded as weighting the Lagrange multipliers 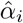 ‘s of positive class so that the minority classification can be improved.

We now apply this idea to LS-SVR. Thus, the classification decision function associated with LS-SVR can be rewritten as follows:

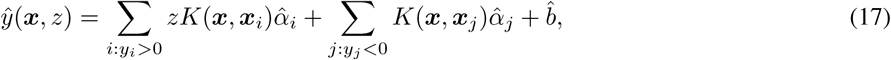

where 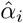 and 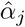 are the Lagrange multipliers associated with positive and negative classes, respectively. The optimal value of a multiplicative weight *z* is obtained by grid search method, which minimizes the quadratic loss function associated with the training data. For simplicity, it is named zLS-SVR. The algorithm of zLS-SVR is given as follows:

1. Train LS-SVR using the training data with the optimal values of hyperparameters.
2. Find the optimal value of multiplicative weight *z*, which minimizes quadratic loss function of the training data.
3. Find the classification decision function *ŷ*(***x***, *z*) for the test data using the optimal value *z*.
4. Classify the test data by using the classification decision function obtained in 3.

### 3.2 weighted LS-SVR

Under the assumption that minority class is considered as positive class, we set the weight 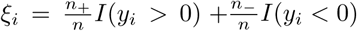 is the number of positive *y*_*i*_’s and *n*_−_ is the number of negative *y*_*i*_’s. Then, the LS-SVR optimization problem in primal weight space is given as follows:

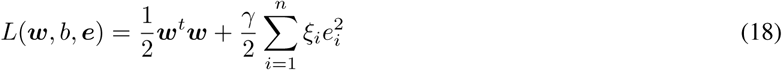

subject to equality constraints

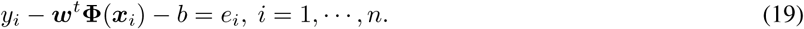

The optimal values of Lagrange multipliers and bias are obtained from the following linear equations

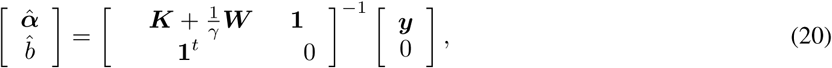

where ***W*** is the diagonal matrix of 1/*ξ*’s. Finally, for a given ***x***_*t*_ the predicted value of response is given as

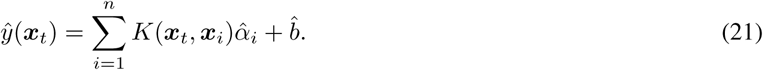

For simplicity, this method is named wLS-SVR.

## 4 Experimental study

In this section we assess the performance of the proposed ensemble classifier through PrOCTOR dataset, which consists of 68 failed drugs for positive class and 708 approved drugs for negative class. The feature vector is divided into ***x*** ∈*R*^17^ (molecular property related) and ***z*** ∈*R*^30^ (target-based property related). We calculate means and standard errors of AUCs for LS-SVR, bLS-SVR, sLS-SVR and random forest (RF) with 100 trees via 5-fold and 10-fold cross validation techniques. We iterate the above procedures 100 times to obtain reliable results. We first investigate how three types of LS-SVR model work for bimodal classification without coping with class-imbalance. Table 1 shows the results for the means and standard errors of 100 AUCs for original dataset. Standard errors are given in parenthesis. Boldfaced values indicate best performance result. As seen from Table 1, stacked LS-SVR model shows the best performance on original dataset.

**Table 1:** AUC results for RF and three types of LS-SVR via 5-fold and 10-fold cross validation techniques without considering class-imbalance

|  | RF | LS-SVR | bLS-SVR | sLS-SVR |
| --- | --- | --- | --- | --- |
| 5-fold CV | 0.6738<br>(0.0017) | 0.6899<br>(0.0017) | 0.6978<br>(0.0014) | **0.7098**<br>(0.0018) |
| 10-fold CV | 0.6785<br>(0.0013) | 0.6894<br>(0.0013) | 0.7016<br>(0.0010) | **0.7157**<br>(0.0011) |

We next investigate how various types of LS-SVR model work for bimodal classification with coping with class-imbalance. We apply synthetic minority over-sampling technique (SMOTE) [16] in advance before training RF, LS-SVR, bLS-SVR and sLS-SVR which are not able to cope with class-imbalance problem. We utilize zLS-SVR and wLS-SVR to overcome imbalance of the data. Table 2 shows the results for the means and standard errors of 100 AUCs. Here, bzLS-SVR, szLS-SVR, bwLS-SVR and swLS-SVR stand for binomial zLS-SVR, stacked zLS-SVR, binomial wLS-SVR and stacked wLS-SVR, respectively. As seen from Table 2, stacked wLS-SVR shows the best performance when considering the class-imbalance. Thus, we recognize that stacked wLS-SVR performs best for bimodal imbalanced PrOCTOR data.

**Table 2:** AUC results for RF and nine types of LS-SVR via 5-fold and 10-fold cross validation techniques for with considering class-imbalance

|  | RF | LS-SVR | bLS-SVR | sLS-SVR |
| --- | --- | --- | --- | --- |
| 5-fold CV | 0.7142<br>(0.0017) | 0.6844<br>(0.0017) | 0.6921<br>(0.0016) | 0.7040<br>(0.0016) |
| 10-fold CV | 0.7172<br>(0.0013) | 0.6888<br>(0.0013) | 0.7010<br>(0.0012) | 0.7119<br>(0.0011) |
|  | zLS-SVR | bzLS-SVR | szLS-SVR |  |
| 5-fold CV | 0.6981<br>(0.0017) | 0.6978<br>(0.0014) | 0.7098<br>(0.0017) |  |
| 10-fold CV | 0.6891<br>(0.0013) | 0.7016<br>(0.0010) | 0.7157<br>(0.0011) |  |
|  | wLS-SVR | bwLS-SVR | swLS-SVR |  |
| 5-fold CV | 0.7111<br>(0.0017) | 0.7130<br>(0.0016) | **0.7204**<br>(0.0016) |  |
| 10-fold CV | 0.7143<br>(0.0070) | 0.7093<br>(0.0072) | **0.7236**<br>(0.0006) |  |

Figure 1 shows AUC results via 5-fold CV for LS-SVR, LS-SVR(SMOTE), zLS-SVR and wLS-SVR. Figure 2 shows AUCs results via 5-fold CV for bLS-SVR, bLS-SVR(SMOTE), bzLS-SVR and bwLS-SVR. Figure 3 shows AUC results via 5-fold CV for sLS-SVR, sLS-SVR(SMOTE), szLS-SVR and swLS-SVR. Figure 4 shows AUC results via 10-fold CV for LS-SVR, LS-SVR(SMOTE), zLS-SVR and wLS-SVR. Figure 5 shows AUCs results via 10-fold CV for bLS-SVR, bLS-SVR(SMOTE), bzLS-SVR and bwLS-SVR. Figure 6 shows AUC results via 10-fold CV for sLS-SVR, sLS-SVR(SMOTE), szLS-SVR and swLS-SVR. Figure 7 shows AUC results via 5-fold and 10-fold CV for RF and RF(SMOTE). From tables and figures we can see that the proposed ensemble classifier shows the best performance for bimodal imbalanced PrOCTOR data.

**Figure 1:**
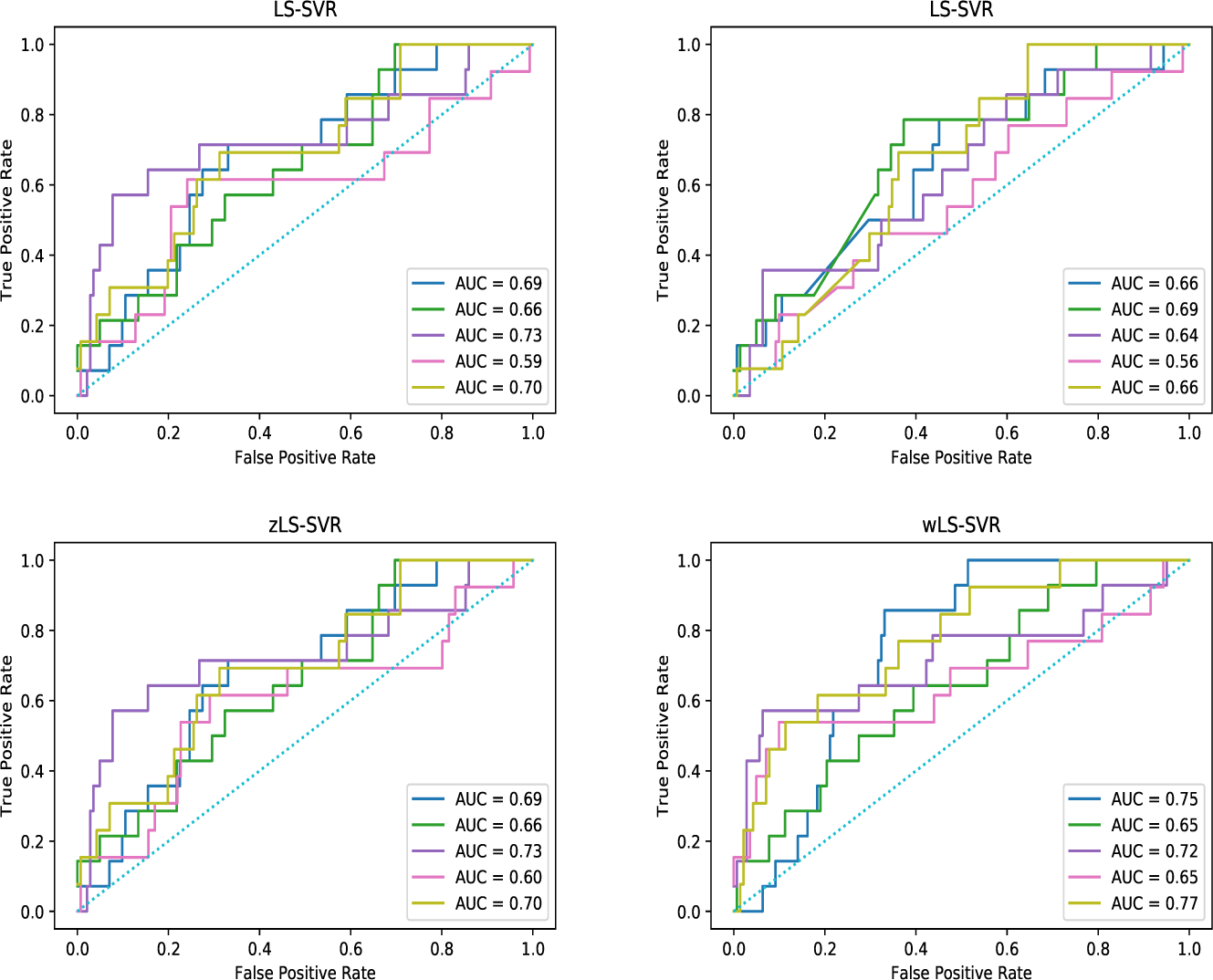
ROC curves for 5-fold CV. LS-SVR (upper left), LS-SVR(SMOTE) (upper right), zLS-SVR (lower left) and wLS-SVR (lower right)

**Figure 2:**
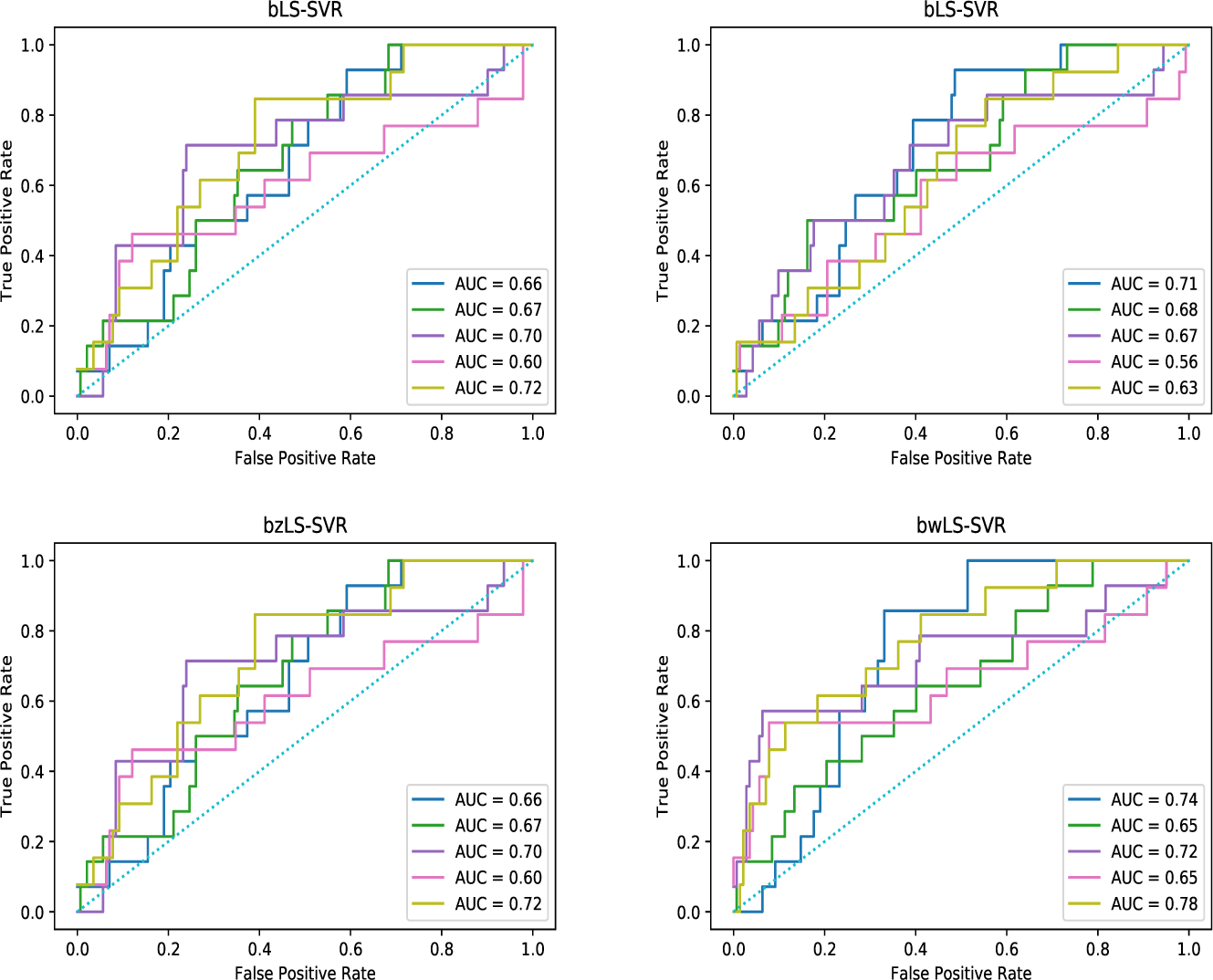
ROC curves for 5-fold CV. bLS-SVR (upper left), bLS-SVR(SMOTE) (upper right), bzLS-SVR (lower left) and bwLS-SVR (lower right)

**Figure 3:**
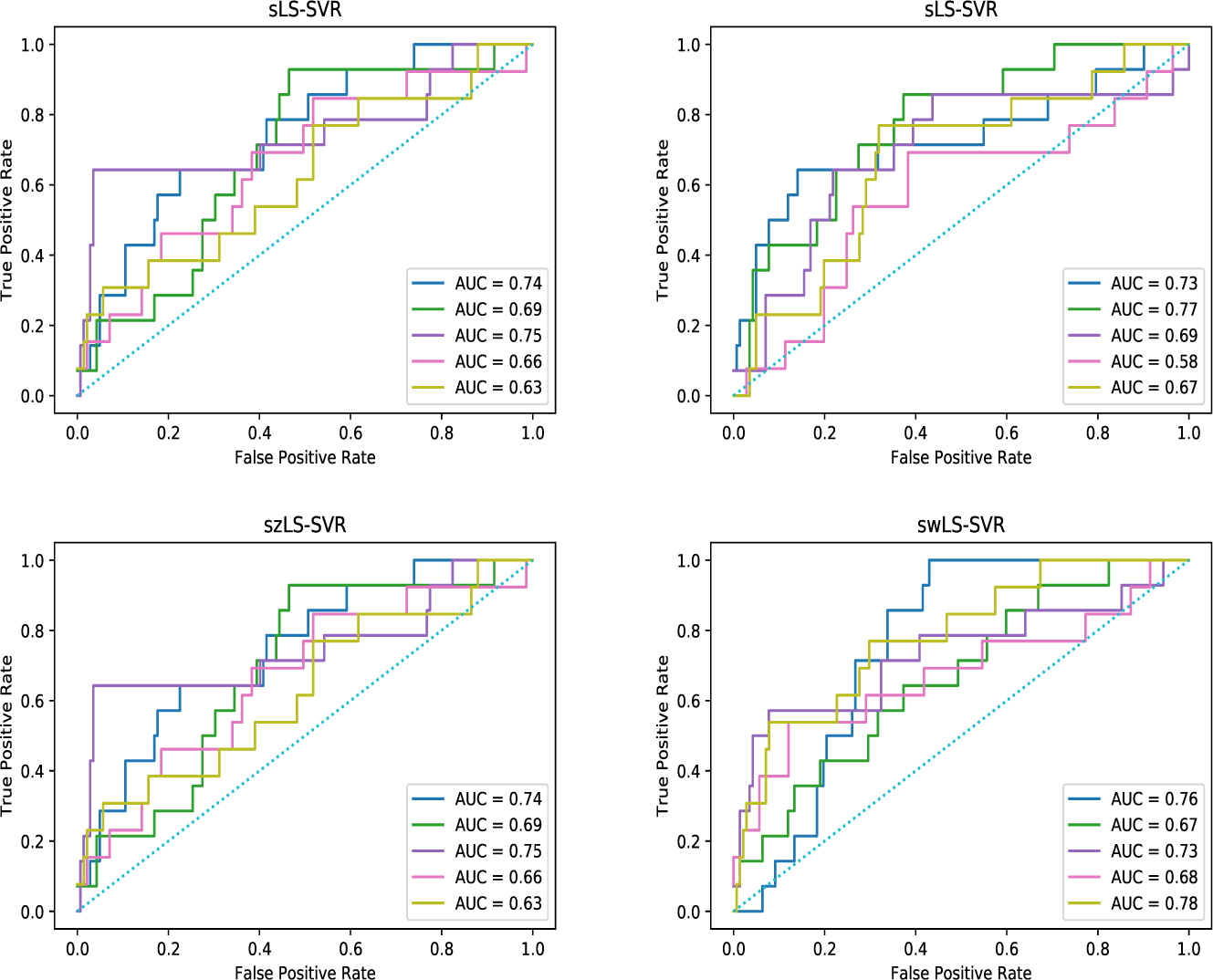
ROC curves for 5-fold CV. sLS-SVR (upper left), sLS-SVR(SMOTE) (upper right), szLS-SVR (lower left) and swLS-SVR (lower right)

**Figure 4:**
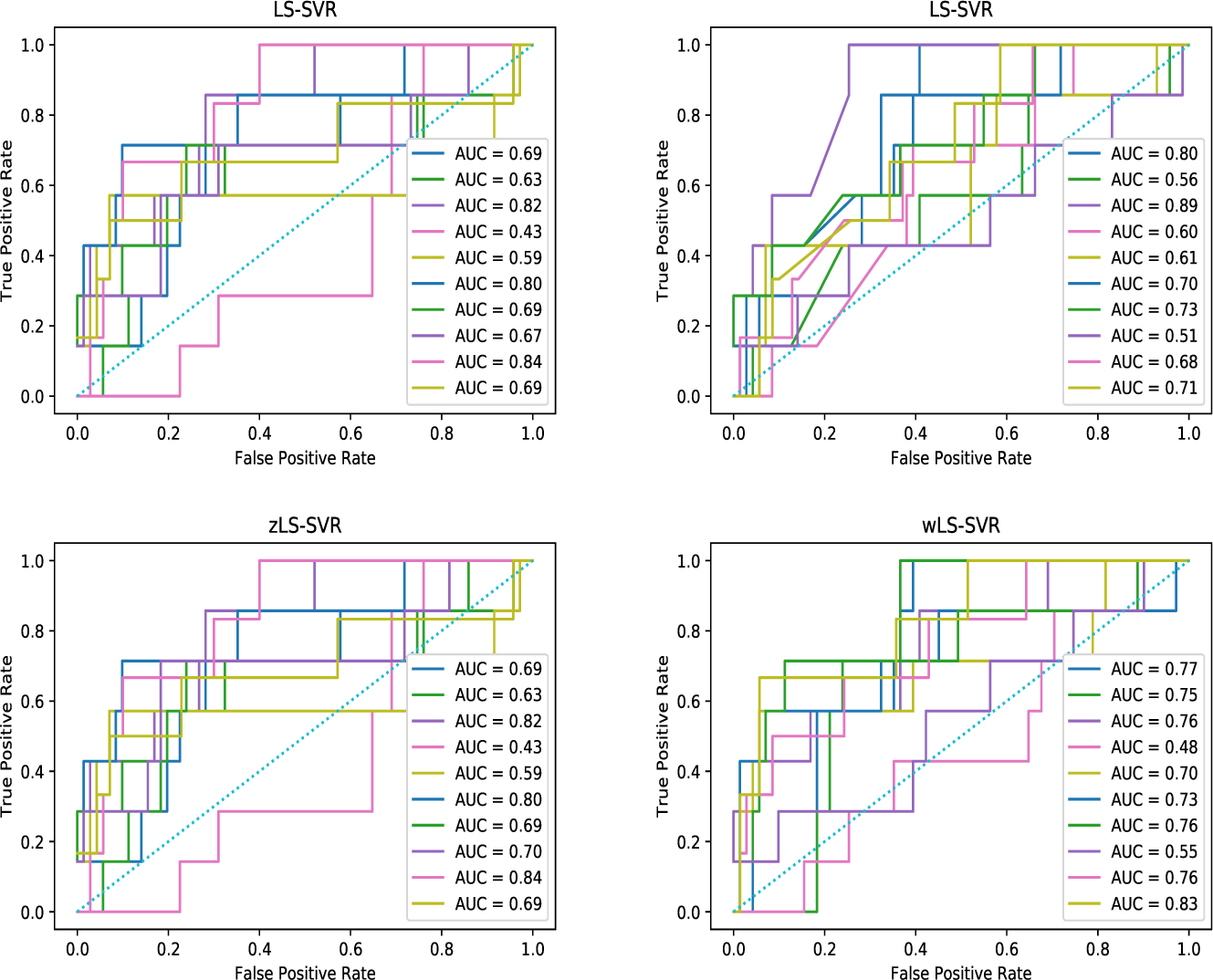
ROC curves for 10-fold CV. LS-SVR (upper left), LS-SVR(SMOTE) (upper right), zLS-SVR (lower left) and wLS-SVR (lower right)

**Figure 5:**
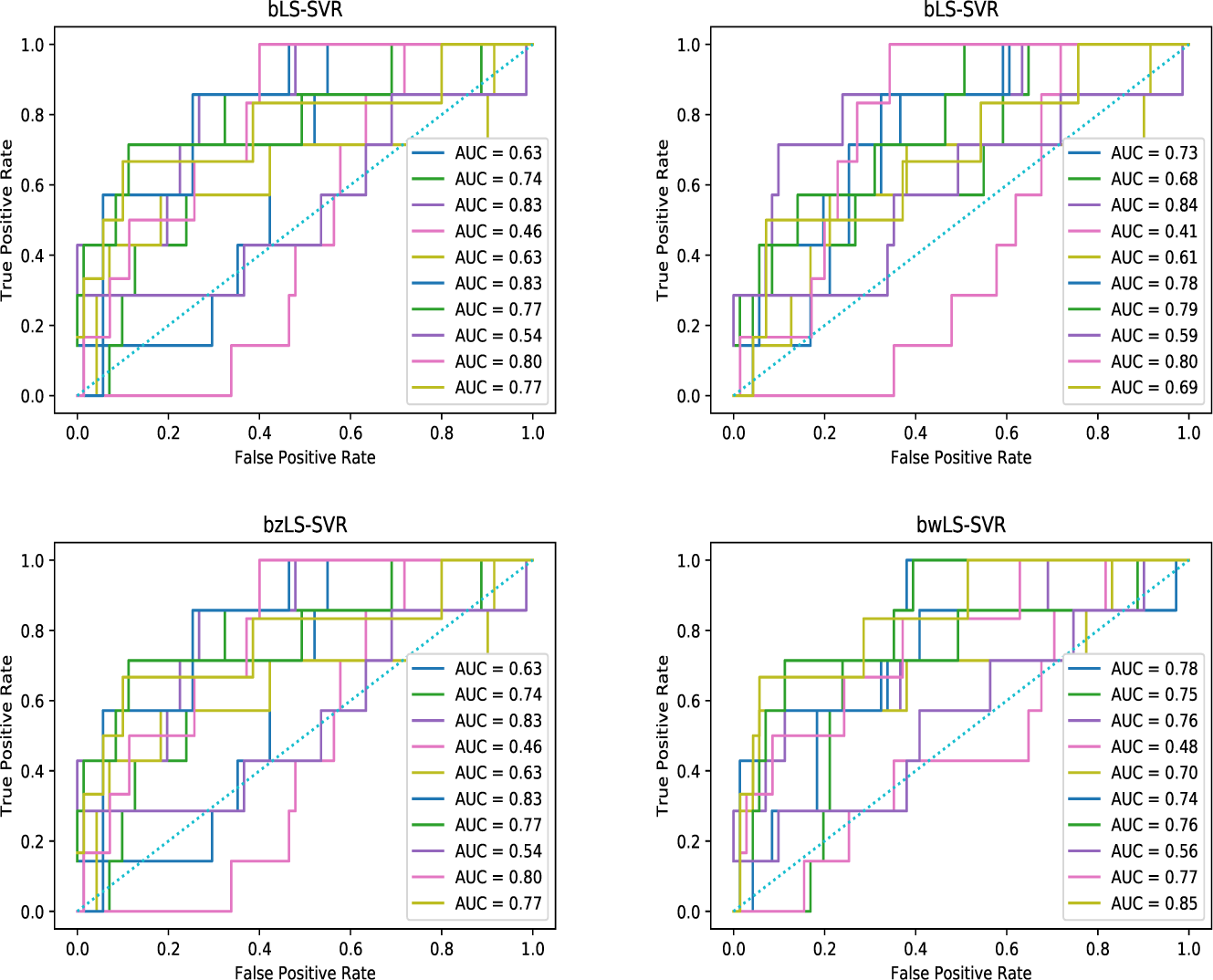
ROC curves for 10-fold CV. bLS-SVR (upper left), bLS-SVR(SMOTE) (upper right), bzLS-SVR (lower left) and bwLS-SVR (lower right)

**Figure 6:**
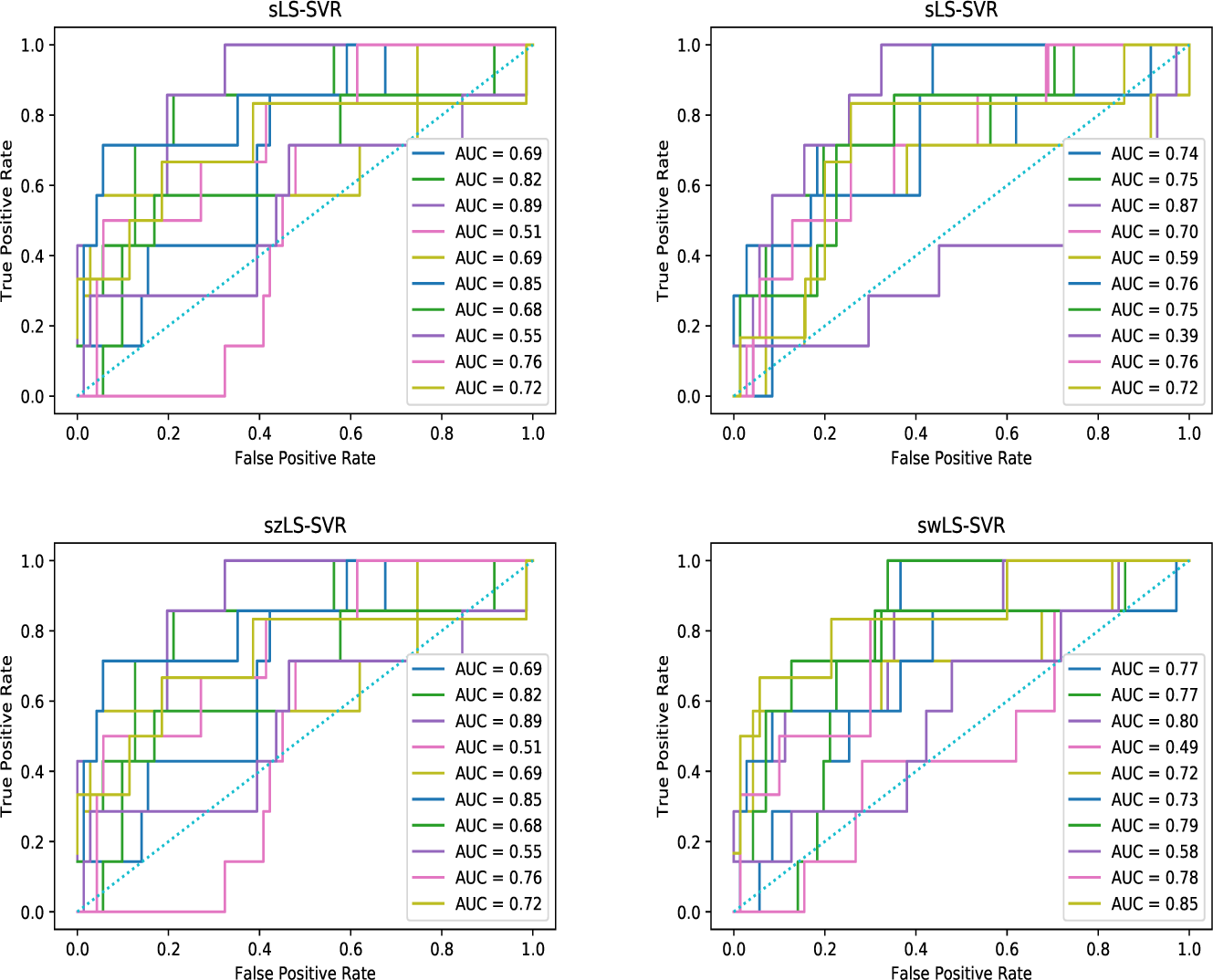
ROC curves for 10-fold CV. sLS-SVR ensemble (upper left), sLS-SVR(SMOTE) (upper right), szLS-SVR (lower left) and swLS-SVR (lower right)

**Figure 7:**
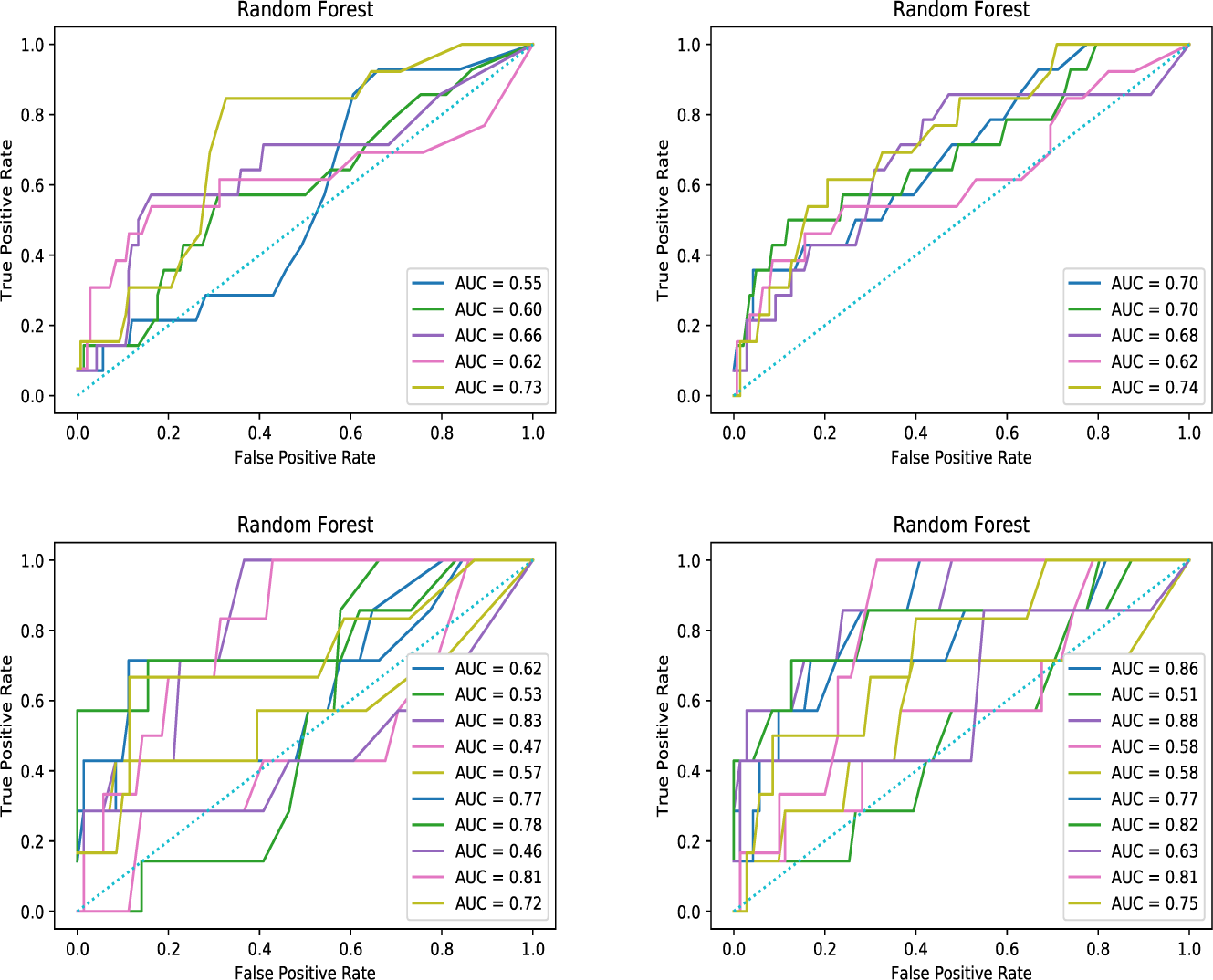
ROC curves for 5-fold & 10-fold CV. RF (5-fold CV, upper left), RF(SMOTE) (5-fold CV, upper right), RF (10-fold CV, lower left) and RF(SMOTE) (10-fold CV, lower right)

## 5 Conclusions

In this paper, we dealt with predicting successes and failures of clinical trials with some variants of LS-SVR model. We found that an ensemble classifier based on weighted LS-SVR provides good results in predicting clinical trial outcomes. In general, the LS-SVR requires knowledge of the related hyperparameters. By the way, the model selection process of LS-SVR makes it possible to obtain the estimates of hyperparameters. Thus, the proposed ensemble classifier appears to be useful in predicting clinical trial outcomes.

To conclude, the proposed ensemble classifier basically has two advantages. One is that this model takes over advantages that SVM works very well for a number of real world problems and overcomes the curse of dimensionality. Thus, the proposed ensemble classifier can be applied easily and effectively to the prediction problem with high dimensional input vector. The other is that this method can predict clinical trial outcomes without knowledge of hyperparameters in advance, because the estimates of hyperparameters are obtained during the model selection process.

## Data Availability

all data referred to in the manuscript are not available.

## Acknowledgements

We thank Olivier Elemento for providing us access to his data. This research was supported by Basic Science Research Program through the National Research Foundation of Korea (NRF) funded by the Ministry of Education (NRF-2018R1D1A1B07042349). This work was supported by “Human Resources Program in Energy Technology” of the Korea Institute of Energy Technology Evaluation and Planning (KETEP), granted financial resource from the Ministry of Trade, Industry & Energy, Republic of Korea (No. 20174030201740). This research was supported by the Bio Medical Technology Development Program of the National Research Foundation (NRF) funded by the Korean government (MSIT) (No. 2019M3E5D4066897).

